# Risk-factors Associated with Non-Vaccination in Gambian Children: A Population-Based Cohort Study

**DOI:** 10.1101/2021.03.19.21253855

**Authors:** Benjamin Young, Golam Sarwar, Ilias Hossain, Grant Mackenzie

## Abstract

**Objective:** We determined the risk-factors associated with children who remain unvaccinated in rural Gambia.

**Methods:** We conducted prospective demographic surveillance and recorded immunisations in real-time in the Basse Health and Demographic Surveillance System. Analysis included residents born between January 1, 2012 and December 31, 2016. Demographic data included age, sex, household members and relationships, migrations, births, deaths, ethnicity, residential location, and birth type. Children were defined as unvaccinated at 10-, 15-, and 24-months of age, if they missed all primary series doses (pentavalent, oral polio and pneumococcal conjugate vaccines), secondary series (1^st^ dose measles and yellow-fever vaccines) or both vaccination series, respectively. Multivariate three-level mixed effects logistic regressions measured the strength of association between risk-factors and being unvaccinated at age 10-, 15-, and 24-months.

**Findings:** 38,090 infants were born during the study period, while 30,832 survived as residents and 1,567 were unvaccinated at age 10 months. Being unvaccinated at 10-months of age was associated with children not residing with their father (adjusted odds ratio [aOR] 1.38, 95% CI 1.22–1.58) or mother (aOR 2.94, 95% CI 1.33–6.46) or both parents (aOR 2.26, 1.60–3.19), whose parents were not the head of household (aOR 1.29 (1.09–1.52), experiencing external in-migration (aOR 2.78, 95% CI 1.52–5.08) and not of Mandinka ethnicity (aOR varied between 1.57 to 1.85 for three other ethnicities).

**Conclusion:** Unimmunised children in rural Gambia are more likely to not live with their parents and have migrated into the area. These results may inform strategies to increase vaccine coverage.

## Introduction

Vaccines have been considered the most successful and cost-effective tool against infectious diseases.^1^ To ensure that vaccines are freely available to every child, the Expanded Programme on Immunisation (EPI) was established nearly 50 years ago by the World Health Organization (WHO).^2^ Vaccination programmes were expected to save more than 20 million lives between 2001 and 2020.^3,4^ However, delayed or missed immunisations in low-and middle-income countries (LMICs) lead to over two million annual vaccine-preventable child deaths.^5–7^

Global vaccination coverage of diphtheria-tetanus-pertussis vaccine’s third dose (DTP3) reached 86% in 2016.^8^ However, high coverage varies geographically and 65% of all unvaccinated children reside in eight African countries.^10^ The Global Vaccine Action Plan 2011-2020, a strategic initiative endorsed by WHO, aimed to increase national vaccination coverage worldwide to over 90% for all national programme vaccines by 2020.^10^ The major under-taking remains in Africa where coverage varies substantially, in West Africa alone, DTP3 coverage varies from 38% in Nigeria to 88% in The Gambia.^11^

From 2009, the Gambian EPI included bacillus Calmette-Guerin (BCG) vaccine, Hepatitis b vaccine (HepB), oral polio vaccine (OPV), DTP, conjugate *Haemophilus influenzae* type b vaccine (Hib), measles vaccine, and Yellow-Fever vaccine.^8^ In 2015, the Gambian EPI added rotavirus vaccine (Rota) and rubella vaccine, a combination vaccine with measles vaccine (MR), while group A meningococcal conjugate vaccine (MenA) was added in 2019 (table 1).^8,9^ Vaccines are administered at Reproductive Child Health (RCH) clinics.

**Table 1.**
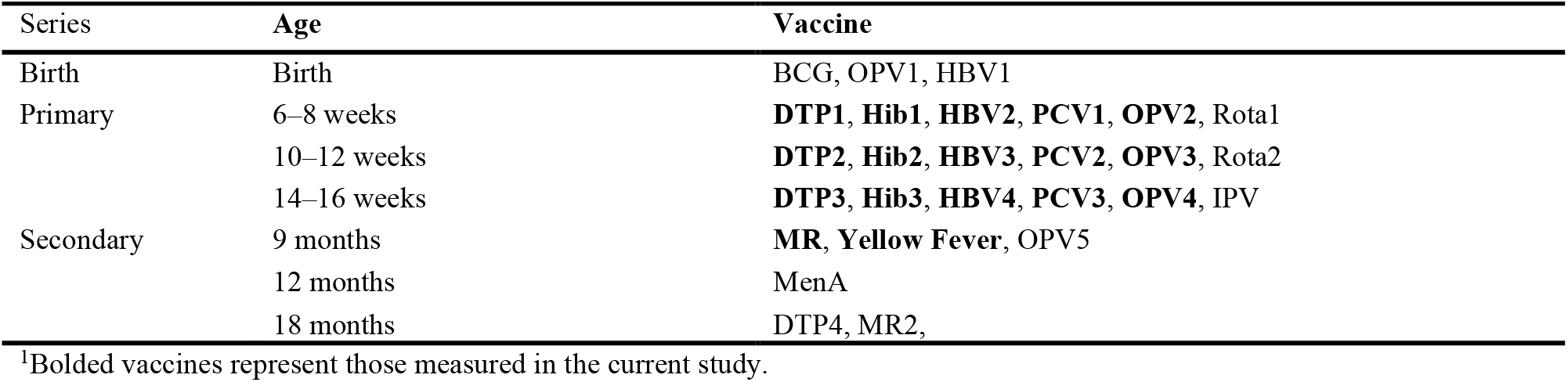
The Gambian vaccination schedule (2020). ^1^.

| Series | Age | Vaccine |
| --- | --- | --- |
| Birth | Birth | BCG, OPV1, HBV1 |
| Primary | 6–8 weeks | **DTP1, Hib1, HBV2, PCV1, OPV2, Rota1** |
|  | 10–12 weeks | **DTP2, Hib2, HBV3, PCV2, OPV3, Rota2** |
|  | 14–16 weeks | **DTP3, Hib3, HBV4, PCV3, OPV4, IPV** |
| Secondary | 9 months | **MR, Yellow Fever, OPV5** |
|  | 12 months | MenA |
|  | 18 months | DTP4, MR2, |

<sup>1</sup>Bolded vaccines represent those measured in the current study.

The Gambian EPI has been a success story and model immunisation programme in sub-Saharan Africa (SSA), with generally high coverage since 1990.^12,13,14^ Part of its success has been due to high public awareness, with accessibility through permanent outreach sites for remote areas and static RCH clinics.^8,13^ To obtain high vaccination coverage, not only is a reliable immunisation programme necessary but usage of the programme by caregivers.^15^ The Gambia, with a well-established vaccination program, represents a unique setting for research into the characteristics of families who don’t utilize a successful programme.

A delay in age appropriate vaccination in The Gambia occurs in two-thirds of children.^12^ Delayed immunisation is associated with the employment status of the mother, birthplace, transportation method, and parental literacy.^2,12^ While researchers often study delayed vaccination,^16^ to the best of our knowledge, we are unaware of African studies reporting risk-factors for being fully unvaccinated.

In Turkey, children of internal migrants or parents with low education were less likely to be vaccinated.^17^ In Nigeria and Liberia, distance to the immunisation facility is an issue in access.^18^ Parental age, the presence of the parents and their marital status have also been associated with ‘under-vaccination’.^19,20^ Reluctance of parents to vaccinate their children is on the rise in high-income countries.^16^ However, vaccine hesitancy is less prevalent in LMICs.^19^

In this study we aimed to identify factors associated with children being fully unvaccinated for the primary series antigens (OPV, PCV and the Pentavalent combination vaccine [DTP-HepB-Hib]) at 10-months of age, the secondary series antigens (1^st^ dose measles and Yellow-Fever) at 15-months of age and for both series of antigens at 24-months of age. We hypothesized that children living further from RCH clinics, are immigrants, and having less educated or absent parents are more likely to be unvaccinated.

## Methods

### Study design and setting

This was a population-based cohort study including residents of the Basse Health and Demographic Surveillance System (BHDSS). The population of the BHDSS was 177,853 in 2014, across 219 settlements. BHDSS residents who were born between January 1, 2012 and December 31, 2016 were included in the study and followed for 24-months.

### Data collection

Demographic surveillance in the BHDSS was conducted through household visits at 4-month intervals.^21^ Data on births, deaths, migrations, household location, education level, head of households and composition of families were collected. Immunisation data was electronically recorded in real time at RCH clinics. A household socio-economic status (SES) survey was conducted in 2012 based on number of assets and livestock. A point system ranked household into wealth quintiles.

### Data analysis

We defined the primary series as three doses each of OPV, PCV and Pentavalent vaccine. At 10-months of age children were categorised as non-vaccinated if they had not received any doses of the primary series vaccines. Children were categorised as vaccinated if they had received one or more doses of any primary series vaccine. The secondary series vaccinations were defined irrespective of the primary series antigens and assessed at 15-months of age, including the first dose of Measles vaccine and Yellow-Fever vaccine. At 24-months of age we categorised children as unvaccinated if they had not received any of their primary or secondary series vaccinations. The birth series vaccination was ignored because we were concerned with the characteristics associated with non-vaccination in infancy. Given our period of follow-up, we ignored vaccines that were introduced after 2012.

The distance between households and RCH clinics was calculated using ArcGIS (Version 10.5) and categorised as distance <0.5 km, ≥0.5–<1 km, ≥1–<2 km, ≥2–<3 km, ≥3–<4 km and ≥4 km. Residents were grouped by type of immigration: none, migration within the BHDSS, internal migration (within The Gambia) or external migration (from outside The Gambia), for each age point of interest. Dates, location of residency and identification of the head of each household were used to determine parental presence for the child and the child’s relationship to the head.

Families were determined based on shared mother IDs and used to determine the birth order of children and type of pregnancy (twin or singleton). The highest educational level achieved by each parent was expressed as: none, basic, secondary, college, Koranic, Madrassa or other. The ethnicity of the child was coded into pre-specified groups: Fula, Serahule, Mandinka and other. Finally, the mother’s age at birth was divided into age groups, age <15 years, ≥15–<20, ≥20–<30, ≥30–<40, and ≥40 years.

Three-level mixed effects (ME) logistic regression, adjusting for nested clustering within families and households were used throughout the analysis.^22^ Univariate analyses assessed the distribution of risk-factors and prevalence of the endpoints using Pearson’s Chi-squared tests and ME logistic regressions. Chi-squared tests produced two-sided p-values and ME logistic regressions produced crude (cOR) with 95% confidence intervals (CI) (table 2 and appendix table 1A). Two multivariate ME logistic regression models were created for each risk factor separately. Model 1 was adjusted including all variables *a priori* to estimate cORs. Model 2 included variables determined as potential confounders in the bivariate analysis, to estimate adjusted odds ratios (aOR). Covariates (table 2) that were weakly associated (p-value <0.20) with the risk factor and vaccination status (at age 10-, 15- or 24-months) and not on the causal pathway were considered potential confounders. Candidate potential confounders were all risk-factors included in table 2. A bivariate analysis determined whether potential confounders were strongly associated (p<0.001) with each other, or collinear. If a controlling variable was collinear with the risk-factor of interest and causing multicollinearity, it was removed from the model. If multicollinearity between strongly associated controlling variables was evident, the variable with the least effect on the model was removed.

**Table 2.** **Descriptive analysis of characteristics of children within the BHDSS and crude and adjusted odds of being unvaccinated^I^ with the primary series of vaccination at 10-months of age.**

| Descriptive variable | Total (Col %)<br>N=30,832 | Unvaccinated (Row %)<br>n=1,567 | Crude OR<br>(95% CI) | p-value<sup>VII</sup> | Adjusted OR<br>(95% CI) | p-value<sup>VII</sup> |
| --- | --- | --- | --- | --- | --- | --- |
| **Sex** |  |  |  |  |  |  |
| Female | 15,076 (48.9) | 775 (5.1) | 1 |  |  |  |
| Male | 15,753 (51.1) | 792 (5.0) | 1.00 (0.89–1.12) | 0.935 |  |  |
| Missing | 3 (0.0) | 0 (0.0) |  |  |  |  |
| **Ethnicity<sup>II</sup>** |  |  |  |  |  |  |
| Mandinka | 6,418 (20.8) | 231 (3.6) | 1 | <0.001 | 1 | <0.001 |
| Fula | 9,836 (31.9) | 525 (5.3) | 1.57 (1.31–1.89) |  | 1.61 (1.33–1.96) |  |
| Serahule | 14,070 (45.6) | 782 (5.6) | 1.64 (1.38–1.95) |  | 1.58 (1.32–1.90) |  |
| Other | 505 (1.6) | 29 (5.7) | 1.85 (1.19–2.89) |  | 1.85 (1.14–3.00) |  |
| Missing | 3 (0.0) | 0 (0.0) |  |  |  |  |
| **Distance to RCH<sup>III</sup>** |  |  |  |  |  |  |
| ≥0 & <0.5 km | 15,113 (49.0) | 737 (4.9) | 1 | 0.261 | 1 | <0.001 |
| ≥0.5 & <1 km | 4,616 (15.0) | 263 (5.7) | 1.21 (1.02–1.44) |  | 1.23 (1.03–1.46) |  |
| ≥1 & <2 km | 4,502 (14.6) | 240 (5.3) | 1.10 (0.92–1.31) |  | 1.16 (0.96–1.40) |  |
| ≥2 & <3 km | 3,051 (9.9) | 151 (4.9) | 1.01 (0.82–1.26) |  | 1.05 (0.84–1.31) |  |
| ≥3 & <4 km | 2,136 (6.9) | 95 (4.4) | 0.92 (0.71–1.18) |  | 0.96 (0.74–1.25) |  |
| ≥4km | 283 (0.9) | 16 (5.7) | 1.11 (0.62–1.98) |  | 1.10 (0.68–2.06) |  |
| Missing | 1,131 (3.7) | 65 (5.7) |  |  |  |  |
| **Migration<sup>IV</sup>** |  |  |  |  |  |  |
| No in-migration | 28,404 (92.1) | 1402 (4.9) | 1 | <0.001 | 1 | <0.001 |
| Within the BHDSS | 551 (1.8) | 43 (7.8) | 1.80 (1.26–2.58) |  | 1.67 (1.15–2.44) |  |
| Internal in-migration | 503 (1.6) | 33 (6.6) | 1.39 (0.91–2.13) |  | 1.22 (0.79–1.89) |  |
| External in-migration | 177 (0.6) | 21 (11.9) | 2.90 (1.61–5.24) |  | 2.60 (1.39–4.87) |  |
| Missing | 1,197 (3.9) | 68 (5.7) |  |  |  |  |
| **Birth order** |  |  |  |  |  |  |
| 1st | 22,223 (72.1) | 1108 (5.0) | 1 | 0.671 |  |  |
| 2nd | 7,485 (24.3) | 362 (4.8) | 0.98 (0.86–1.12) |  |  |  |
| 3rd | 601 (1.9) | 36 (6.0) | 1.25 (0.85–1.83) |  |  |  |
| 4th or higher | 30 (0.1) | 2 (6.7) | 1.24 (0.23–6.81) |  |  |  |
| Missing | 493 (1.6) | 59 (12.0) |  |  |  |  |
| **Pregnancy type** |  |  |  |  |  |  |
| Singleton | 29,324 (95.1) | 1452 (5.0) | 1 |  |  |  |
| Twins | 1,015 (3.3) | 56 (5.5) | 1.09 (0.75–1.59) | 0.647 |  |  |
| Missing | 493 (1.6) | 59 (12.0) |  |  |  |  |
| **Head of house<sup>V</sup>** |  |  |  |  |  |  |
| Was a parent | 6,020 (19.5) | 247 (4.1) | 1 |  | 1 |  |
| Was not a parent | 24,471 (79.4) | 1298 (5.3) | 1.31 (1.12–1.53) | <0.001 | 1.29 (1.09–1.52) | <0.001 |
| Missing | 341 (1.1) | 22 (6.5) |  |  |  |  |
| **Mothers age at birth** |  |  |  |  |  |  |
| <15 | 178 (0.6) | 10 (5.6) | 1 | 0.262 |  |  |
| ≥15 & <20 | 3,763 (12.2) | 194 (5.2) | 0.93 (0.46–1.89) |  |  |  |
| ≥20 & <30 | 16,244 (52.7) | 838 (5.2) | 0.93 (0.47–1.86) |  |  |  |
| ≥30 & <40 | 8,799 (28.5) | 399 (4.5) | 0.80 (0.40–1.61) |  |  |  |
| ≥40 | 1,338 (4.3) | 67 (5.0) | 0.89 (0.42–1.87) |  |  |  |
| Missing | 510 (1.7) | 59 (11.6) |  |  |  |  |
| **Presence of parents<sup>VI</sup>** |  |  |  |  |  |  |
| Both present | 13,938 (45.2) | 574 (4.1) | 1 | <0.001 | 1 | <0.001 |
| Mother present | 15,605 (50.6) | 867 (5.6) | 1.40 (1.25–1.58) |  | 1.38 (1.22–1.58) |  |
| Father present | 155 (0.5) | 12 (7.7) | 2.78 (1.32–5.88) |  | 2.93 (1.33–6.46) |  |
| Neither present | 1,134 (3.7) | 114 (10.1) | 2.26 (1.62–3.15) |  | 2.26 (1.60–3.19) |  |
| **Wealth quintile** |  |  |  |  |  |  |
| 1st quintile | 2,851 (9.2) | 138 (4.8) | 1 | 0.901 |  |  |
| 2nd quintile | 3,258 (10.6) | 162 (5.0) | 1.04 (0.79–1.36) |  |  |  |
| 3rd quintile | 3,506 (11.4) | 196 (5.6) | 1.14 (0.88–1.47) |  |  |  |
| 4th quintile | 5,467 (17.7) | 284 (5.2) | 1.08 (0.85–1.37) |  |  |  |
| 5th quintile | 6,491 (21.1) | 335 (5.2) | 1.06 (0.84–1.33) |  |  |  |
| Missing | 9,259 (30.0) | 452 (4.9) |  |  |  |  |

<sup>I</sup>Children were defined as unvaccinated if they had not received any primary series vaccinations (OPV, PCV, and Pentavalent) by 10-months of age.
<sup>II</sup>Controlled for sex, mothers age at birth, distance from health centre, presence of parents, immigration and headship
<sup>III</sup>Controlled for sex, mother's age at birth, ethnicity, presence of parents, immigration and headship
<sup>IV</sup>Controlled for sex, mother's age at birth, ethnicity, distance from health centre, presence of parents and headship
<sup>V</sup>Controlled for sex, mother's age at birth, ethnicity, immigration and distance from health centre
<sup>VI</sup>Controlled for sex, mother's age at birth, ethnicity, distance from health centre and immigration
<sup>VII</sup>P-values obtained using Wald test

Cox proportional hazards models were used to evaluate the association between vaccination status and all-cause mortality. The association between vaccination status at age 10-months and the incidence of death between 10- and 24-months of age was expressed as hazard rate ratios (HR); the same was done for vaccination status at age 15-months and incidence of death between 15- and 24-months of age. Analyses excluded deaths before 10- and 15-months of age respectively to remove the bias of a likely association with non-vaccination. Potential confounders were included using similar methods as previously described. The proportional hazards assumption was assessed graphically and tested using Schoenfeld residuals.

### Missing data and sensitivity analysis

Children without a 14-digit ID were not fully enumerated and considered non-residents of the BHDSS. Non-residents were excluded from the analysis. Covariates with significant missing data (missingness ≥10%) were removed from the primary analysis. Sensitivity analyses, using logistic regressions, were performed to check whether data were missing at random (MAR). Strong evidence of differentially missing data were considered not missing at random (NMAR) and discussed in the limitations.

Data were cleaned, validated, and analysed using Stata (Version 15.1). Ethical approval was obtained from the LSHTM Ethics Committee before the data were obtained and analysis begun.

## Results

There were 30,832 resident children recorded at 10-months of age in the BHDSS between January 1, 2012 and December 31, 2016 as well as 30,561 and 30,120 residents at the 15- and 24-month age marks.

### Descriptive analysis

Of all resident children at 10-months of age 5.1% (1,567/30,832) were unvaccinated with the primary series. At 15-months of age 22.5% (6,886/30,561) of children were unvaccinated with the secondary series and 3.3% (981/30,210) were unvaccinated at 24-months of age with both primary and secondary series.

The distributions of risk-factors at 10-months of age are shown in table 2. Most children (49.0%) lived <0.5 km from an RCH clinic, and only 283 (0.9%) resided ≥4 km away. Only 1,170 (3.8%) children had experienced any type of immigration before 10-months of age. Less than half of the children, (45.2%) shared a residence with both their parents at age 10-months, most children, (50.6%), lived solely with their mother. In terms of SES, 9,258 (30.0%) of the children lived in a household missing a wealth score.

The distributions of risk-factors for the 15- and 24-month vaccination status analysis are found appendix table 1A and followed similar patterns as the primary analysis.

Children were grouped into 22,152 families within 8,315 households, (median of four families per household and two children per family), 493 (1.6%) children were missing family data. The geographic locations of households and RCH facilities are represented in figure 1.

**Figure 1.**
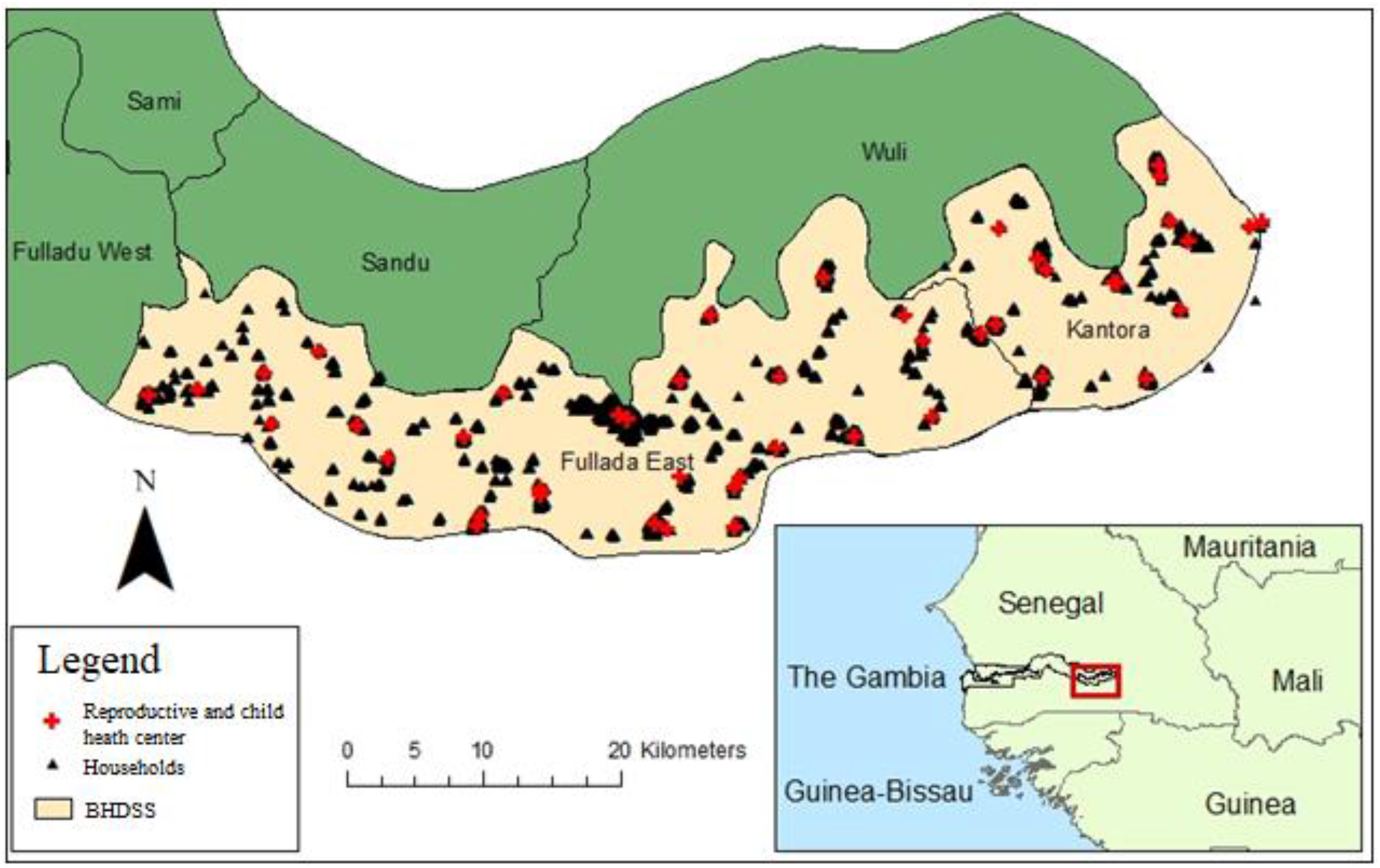
**Map of household locations and Reproductive and Child Health clinics in the Basse Health and Demographic Surveillance System in eastern Gambia.**

### Univariate analysis

#### Primary analysis at 10-months of age

At 10-months of age there was no evidence of a relationship between distance from an RCH clinic and children who had missed their primary vaccines (p=0.261) (table 2). There was a very strong association (p<0.001) between the presence of parents and vaccination status. As well, if parents were not the household head, the child had 31% higher odds of missing their primary series (cOR 1.31, 95% CI 1.12–1.53, p<0.001). Children who had externally in-migrated before age 10-months had nearly three times higher odds of missing their primary series compared to non-migrants (cOR 2.90, 95% CI 1.61–5.24, p<0.001). The Mandinka ethnicity had the lowest proportions of unvaccinated children (3.6%) and Serahule children had 64% higher odds of being unvaccinated (cOR 1.64, 95% CI 1.38–1.95, p<0.001). There was no association (p=0.901) between the wealth quintile of a household and the child’s vaccination status. Sex, pregnancy type, birth order and mother’s age at birth were not associated with vaccination status at 10-months of age (p=0.935, p=0.647, p=0.671 and p=0.262, respectively).

#### Secondary analysis at 15- and 24-months of age

There were similar associative patterns observed at age 15- and 24-month vaccination status, compared to age 10-months status. An important difference was the strong evidence for an association between birth order and age 15- and 24-month vaccination status (p<0.001 and p=0.016, respectively). In both age groups third-born children had the largest increase in odds for being unvaccinated compared to first-borns, (cOR 2.39, 95% CI 1.94–2.95 and cOR 1.80, 95% CI 1.15– 2.80). A full description for the secondary univariate analysis can be found in the appendix and table 1A.

### Multivariate analysis

#### Primary analysis at 10-months of age

Based on univariate analyses and our hypotheses, the child’s ethnicity, residential distance from the RCH clinic, relationship to the head of household, parental presence and immigration status were considered important risk-factors and assessed in multivariate analysis (table 2). The child’s sex and mother’s age at birth were considered *a priori* confounders. The covariates considered as potential confounders and included in each of the fully adjusted models are listed in table 2.

After adjusting for confounders and clustering there was strong evidence (p<0.001) that Fula and Serahule children had higher odds of being unvaccinated compared to Mandinka children (aOR 1.64, 95% CI 1.30–2.08 and aOR 1.68, 95% CI 1.35–2.09, respectively). There was weak evidence for increased odds of non-vaccination among children residing between ≥0.5–<1 km from an RCH compared to <0.5 km (aOR 1.25, 95% CI 1.01–1.46). There was no evidence for a difference in the odds of unvaccinated children residing other distances from an RCH clinic as all CI’s contained unity (table 2).

After adjusting for confounders, children residing with a single mother had odds of being unvaccinated 38% higher than those residing with both parents, if they resided solely with the father the odds increased by 194% (aOR 1.38, 95% CI 1.22–1.58 and aOR 2.94, 95% CI 1.33–6.46).

In the adjusted model, there remained strong evidence (p<0.001) for a negative effect on vaccination status associated with external in-migration compared to no migration despite signs of confounding reducing the effect (aOR 2.60, 95% CI 1.39–4.87). Compared to non-migrants, there was no evidence (p=0.380) that children who internally in-migrated, and strong evidence (p=0.008) for those who migrated within the BHDSS, of an increase in odds of being unvaccinated (table 2).

Despite evidence of positive confounding in the adjusted model, a reduction in the effect estimate towards the null, there remained strong evidence (p<0.001) that a child without a parent as the head of the household had 29% higher odds of being unvaccinated compared to a child with either parent holding the position (aOR 1.29, 95% CI 1.09–1.52).

There was evidence of multicollinearity (>15% change in log standard errors) amongst controlling covariates when the wealth quintile risk-factor was included in all fully adjusted models. As a controlling variable the wealth quintile did not indicate signs of important confounding when added to any adjusted models, save for when estimating the effect of a single father compared to baseline, removing evidence of an effect (aOR 1.73, 95% CI 0.57–5.23). However, when included in the model, strong signs of multicollinearity remained between covariates.

#### Secondary analysis at 15- and 24-months of age

In the multivariate analysis there were similar associative patterns between risk-factors and unvaccinated status at age 15- and 24-months as there were at age 10-months (appendix and table 2A).

### Death analysis

The Cox proportional hazards model showed no evidence of an association (p=0.338) between vaccination status at age 10-months and incidence of mortality between 10- and 24-months of age (aHR 1.25, 95% CI 0.79–2.00). There was strong evidence (p<0.001) of an association between 15-month vaccination status and mortality between age 15- and 24-months (aHR 1.81, 95% CI 1.32– 2.48). Both 10- and 15-month vaccination status survival models satisfied the proportional hazards assumption (appendix figure 1A and figure 2A).

### Missing data and sensitivity analysis

5,603 (13.7%) children were not fully enumerated and therefore not confirmed resident within the BHDSS and excluded. Sensitivity analysis found the unenumerated children NMAR and differentially missing on both outcome and exposure variables. Unenumerated children had higher odds of being unvaccinated compared to fully enumerated children (cOR 2.53, 95% CI 2.31–2.77). As well, strong evidence (p<0.001) indicates the excluded subgroup were also born to a higher proportion of mothers less than 15-years old, 1.7% vs. 0.6%.

In the primary dataset there were 9,259 (30.0%) children with missing household wealth quintile. Wealth data were MAR across the outcome with no association between missingness and vaccination status (p=0.293). Education level was missing for 62% of mothers and 83% of fathers. Children with missing mother and father education data had increased odds of non-vaccination (cOR 1.18, 95% CI 1.06–1.31 and cOR 1.41, 95% CI 1.21–1.63, respectively).

## Discussion

We found that 1,567 (5.1%) resident children at age 10-months had missed all their primary series vaccines. There was strong evidence that children who (1) had at least one parent missing, (2) had experienced external in-migration, (3) whose parents were not the head of household, or (4) were not Mandinka, had increased odds of being unvaccinated with their primary series compared to their counterparts.

The literature suggests that distance to an immunisation clinic is a major cause of under-vaccination. ^20,23,24^ Our findings showed that distance to an RCH clinic, a spatial issue, was not associated with non-vaccination. Large distances impede immunisation uptake^23^ but distance may not associate with non-vaccination in settings where shorter distances are involved.

Literature also suggests increasing birth order is associated with delayed immunisation,^2^ whereas our research found no association with non-vaccination. Increasing birth order may derive from a temporal issue (i.e. busier parents) and not impede awareness or drive negligence towards receiving immunisations. It appears in The Gambia that factors related to spatial or temporal issues may associate more with delayed or missed immunisations whereas those related to programme awareness, migration, or levels of social care for vulnerable children may associate more with complete non-vaccination.

A Turkish study found an association between non-vaccination and in-migrant children, consistent with our findings in terms of external in-migration.^17^ This likely arises from a complex set of awareness or accessibility issues; emigrating from areas without established immunisation programmes, lack of knowledge of the local immunisation programme or missing immunisation records.^17^ Our analysis found no evidence of a negative effect from internal in-migration or migration within the BHDSS. This may diverge from the literature as immunisation coverage across The Gambia is quite stable and consistently recorded, Gambian children in-migrating may have had equal access to immunisations prior to departing their previous residential location.

Mandinka children had a lower proportion of non-vaccinated children, which is consistent with previous literature.^8,25^ Ethnicity as a risk factor may derive from education and attitudinal issues and rural Serahules and Fula may be more doubtful of immunisation programmes.

Children living with one parent had higher odds of being unvaccinated which is consistent with current literature.^26,27^ Single parenthood may combine temporal and awareness issues, where a caregiver isn’t available to take the child for immunisation, vaccines may be a lower priority for a busier parent,^15^ or with only one caregiver, they are less likely to be aware of immunisation programmes.

The lack of an association between the primary series immunisation status at 10-months and mortality between 10- and 24-months is understandable. Literature shows high-immunisation coverage in the first year of life over the past 10-years in The Gambia and unvaccinated children benefit from herd immunity and related reduction in vaccine preventable deaths.^8^ The increased risk of mortality between 15- and 24-months of age in children who remained unvaccinated with their secondary series may also be consistent with literature, that suggests measles vaccine may have non-specific beneficial effects.^28^ Researchers found a reduction in all-cause mortality of 30–86% for children who received the standard measles immunisations.^28^

The size of our study meant our rare primary outcome was observed with sufficient frequency, allowing consideration of multiple covariates. Our study had over 95% power to detect a 20% increase in odds, giving a very low probability of type 1 error. The systematic enumeration in the BHDSS reduced selection bias via sampling error. As this was a population-based study estimates of effect are representative of the population. Demographic data collected every 4 months reduces measurement error in frequently changing variables and provides temporality, removing the possibility of reverse causality. Our real-time electronic recording of immunisations eliminates recall and information bias. Vaccination coverage studies often rely on the memory of parents^15^ or easily lost vaccination cards,^26^ to determine a child’s vaccinations status.

Our study is limited by the variables collected by the BHDSS. Unmeasured risk-factors could result in residual or uncontrolled confounding. Factors such as perception of vaccine safety or pregnancy outside of marriage, may independently explain some of our observed effects. Over a quarter of the children had no SES data. The sensitivity analysis found the missingness to be non-differential, however this loss of power may have caused a type II error in our results, with potentially increased probability of false negative findings. The household SES data was not included in the final model due to strong evidence of multicollinearity between the available SES data and covariates. The mechanism of associations and collinearity between the controlling covariates and SES is not well understood. It seems plausible, due to the lack of observed confounding in the adjusted model when SES is included, the effect associated with SES may partially be controlled through collinear variables in the analysis. Parental education level was intended to be included in the analysis and deemed a likely confounder.^17^ However the data were largely missing (>65%) and missingness at this magnitude meant the available data would not give reliable information. This remains a limitation to the study and parents with varying education levels may explain some of the observed effect on children being unvaccinated at 10-months of age. Some selection bias is likely for the subgroup of unenumerated children (13.7%) that were excluded. There was strong evidence that this subgroup had higher odds of being unvaccinated at age 10-months and their removal from the analysis could bias the results in either direction. Children who are unvaccinated may be more likely to be sick or unhealthy, and children who died before 10-months of age were excluded from the analysis. Our analysis does not relate to such children. Another limitation was our definition of in-migration only applying after a child’s birth. This may differentially misclassify immigration biasing the effect toward the null if the negative effect on non-vaccination lingers on immigrant parents despite the child being a native resident.

Broad assumptions were made when categorising the presence of parents. Varying rates of non-vaccination for children grouped within parameters could produce bias or residual confounding. The unknown status of the absent parent(s) (e.g. deceased, remarried, or immigrant worker) may have unmeasured effects on the odds of vaccination, biasing the estimated effect towards the null.

Our findings suggests that the factors which lead to unvaccinated children in The Gambia are multifaceted. It is unlikely that the limitations of the study would completely remove the observed effects that these risk-factors have on being unvaccinated. The external validity of some risk-factors may be limited by the unique characteristics of The Gambia, such as ethnicity where demographics and social structures may vary by country. However, sociological factors such as parental presence, authorities within a household and immigration remain generalisable to other SSA countries. National vaccination programme officials seeking to optimise their vaccination programmes should consider our findings, targeting the aforementioned risk-factors to reduce rates of non-vaccination. Future studies should consider vaccine hesitancy, to better understand this trend in The Gambia.

## Data Availability

Data may be available per requests.

## Appendix

### Results

#### Secondary univariate analysis

The secondary univariate analysis at 15- and 24-months of age generally followed the same patterns of association as with vaccination status at age 10-months (table 1A and table 2A). There was strong evidence of an association between unvaccinated children and birth order at 15- and 24-months of age (p-value <0.001 and p-value=0.016, respectively). Third born children had 2.39 and 1.80 higher odds at age 15- and 24-months compared to first born children (cOR 2.39, 95% CI 1.94–2.95 7 and cOR 1.80, 95% CI 1.15 - 2.80).

There was no evidence in the univariate analysis of an association between distance of a residence from the RCH and the vaccination status at both 15- and 24-months of age (p=0.108 and p=0.435, respectively). There was no evidence for an association between the pregnancy type and the vaccination status at 15- and 24-months of age (p=0.194 and p=0.681, respectively). There was strong evidence for an association between the immigration status of a child at both 15- and 24- months of age and their vaccination status (p<0.001 and p=0.002). At both 15- and 24-months of age, children who experienced external in-migration had 70% and 175% higher odd of being unvaccinated, respectively, compared to children who had not experienced any in-migration (cOR 1.70, 95% CI 1.16–2.48 and cOR 2.75, 95% CI 1.55–4.87, respectively).

At 15- and 24-months of age there was strong evidence of an association between the ethnicity of the child and their respective vaccination status (p-value <0.001 & p-value <0.001). Mandinka ethnicity had the lowest proportion of unvaccinated children at age 15-months (16.9 %) and age 24-months (2.1%). Compared to Mandinka children, at 15-months of age Fula children had the largest increase in odds of non-vaccination, (cOR 1.74, 95% CI 1.65–1.93), at 24-months of age Fula and Serahule children had similar increased odds of non-vaccination compared to Mandinka children (cOR 1.64, 95% CI 1.30–2.08 and cOR 1.68, 95% CI 1.35–2.09, respectively).

Children whose parent(s) was not the head of household had higher odds of being unvaccinated at 15- and 24-months of age (cOR 1.11, 95% CI 1.01–1.21 and cOR 1.32, 95% CI 1.08–1.61, respectively). There was no evidence of an association between the mother’s age at birth and the vaccination status of the child at both 15- and 24-month age points of interest (p=0.124 and p=0.570, respectively). There was a strong association between the presence of parents and a child’s vaccination status at 15- and 24-months of age (p<0.001 and p<0.001, respectively). The crude analysis showed the greatest effect when comparing parameters to both parents being present in children with neither parent present whose odds of non-vaccination tripled and doubled, respectively (cOR 2.98, 95% CI 2.45–3.62 & cOR 1.99, 95% CI 1.38–2.87). There was no evidence of an association between the wealth quintile of the household and the vaccination status of the child at 15- and 24-months of age (p=0.669 and p=0.568, respectively).

**Table 1A.** **Descriptive analysis of characteristics of children within the BHDSS and the crude and adjusted odds of being unvaccinated with the secondary vaccination series at 15-months of age.**

| Descriptive variable | Total (Col %)<br>N=30,561 | Unvaccinated (Row %)<br>n=6,886 | Crude OR<br>(95% CI) | p-value<sup>VII</sup> | Adjusted OR<br>(95% CI) | p-value<sup>VII</sup> |
| --- | --- | --- | --- | --- | --- | --- |
| **Sex** |  |  |  |  |  |  |
| Female | 14,943 (48.9) | 3,365 (22.5) | 1 | 0.124 |  |  |
| Male | 15,615 (51.1) | 3,521 (22.5) | 1.01 (0.95–1.07) |  |  |  |
| Missing | 3 (0.0) | 0 (0.0) |  |  |  |  |
| **Ethnicity<sup>II</sup>** |  |  |  |  |  |  |
| Mandinka | 6,365 (20.8) | 1,075 (16.9) | 1 | <0.001 | 1 | <0.001 |
| Fula | 9,717 (31.8) | 2,424 (24.9) | 1.74 (1.65–1.93) |  | 1.81 (1.62–2.02) |  |
| Serahule | 13,986 (45.8) | 3,286 (23.5) | 1.56 (1.41–1.72) |  | 1.56 (1.41–1.74) |  |
| Other | 490 (1.6) | 101 (20.6) | 1.29 (0.98–1.71) |  | 1.26 (0.93–1.70) |  |
| Missing | 3 (0.0) | 0 (0.0) |  |  |  |  |
| **Distance to RCH<sup>III</sup>** |  |  |  |  |  |  |
| ≥0 & <0.5 km | 14,985 (49.0) | 3,277 (21.9) | 1 | 0.108 | 1 | <0.001 |
| ≥0.5 & <1 km | 4,565 (14.9) | 1,072 (23.5) | 1.14 (0.94–1.17) |  | 1.12 (1.00–1.26) |  |
| ≥1 & <2 km | 4,473 (14.6) | 1,019 (22.8) | 1.05 (0.94–1.17) |  | 1.01 (0.90–1.13) |  |
| ≥2 & <3 km | 3,012 (9.9) | 694 (23.0) | 1.07 (0.94–1.23) |  | 1.11 (0.97–1.27) |  |
| ≥3 & <4 km | 2,109 (6.9) | 475 (22.5) | 1.09 (0.94–1.26) |  | 1.12 (0.97–1.29) |  |
| ≥4 km | 274 (0.9) | 55 (20.1) | 0.82 (0.52–1.31) |  | 0.84 (0.54–1.32) |  |
| Missing | 1,143 (3.7) | 294 (25.7) |  |  |  |  |
| **Migration<sup>IV</sup>** |  |  |  |  |  |  |
| No in-migration | 27,736 (90.8) | 6,145 (22.2) | 1 | <0.001 | 1 | <0.001 |
| Within the BHDSS | 767 (2.5) | 219 (28.6) | 1.52 (1.25–1.84) |  | 1.39 (1.13–1.72) |  |
| Internal in-migration | 635 (2.1) | 159 (25.0) | 1.19 (0.94–1.51) |  | 1.19 (0.93–1.51) |  |
| External in-migration | 231 (0.8) | 78 (33.8) | 1.70 (1.16–2.48) |  | 1.75 (1.12–2.59) |  |
| Missing | 1,192 (3.9) | 285 (23.9) |  |  |  |  |
| **Birth order** |  |  |  |  |  |  |
| 1st | 21,996 (72.0) | 4,626 (21.0) | 1 | <0.001 |  |  |
| 2nd | 7,420 (24.3) | 1,822 (24.6) | 1.30 (1.21–1.40) |  |  |  |
| 3rd | 598 (2.0) | 208 (34.8) | 2.39 (1.94–2.95) |  |  |  |
| 4th or higher | 30 (0.1) | 9 (30.0) | 1.70 (0.65–4.42) |  |  |  |
| Missing | 517 (1.7) | 221 (42.7) |  |  |  |  |
| **Pregnancy type** |  |  |  |  |  |  |
| Singleton | 29,035 (95.0) | 6,439 (22.2) | 1 | 0.194 |  |  |
| Twins | 1,009 (3.3) | 226 (22.4) | 0.99 (0.79–1.24) |  |  |  |
| Missing | 517 (1.7) | 221 (42.7) |  |  |  |  |
| **Head of house<sup>V</sup>** |  |  |  |  |  |  |
| Was a parent | 6,044 (19.8) | 1,279 (21.2) | 1 | 0.044 | 1 | 0.076 |
| Was not a parent | 24,181 (79.1) | 5,515 (22.8) | 1.11 (1.01–1.21) |  | 1.09 (0.99–1.19) |  |
| Missing | 336 (1.1) | 92 (27.4) |  |  |  |  |
| **Mothers age at birth** |  |  |  |  |  |  |
| <15 | 176 (0.6) | 36 (20.5) | 1 | 0.124 |  |  |
| ≥15 & <20 | 3,720 (12.2) | 789 (21.2) | 1.11 (0.73–1.70) |  |  |  |
| ≥20 & <30 | 16,066 (52.6) | 3,658 (22.8) | 1.23 (0.81–1.85) |  |  |  |
| ≥30 & <40 | 8,740 (28.6) | 1,892 (21.6) | 1.12 (0.72–1.69) |  |  |  |
| ≥40 | 1,325 (4.3) | 286 (21.6) | 1.10 (0.71–1.70) |  |  |  |
| Missing | 534 (1.7) | 225 (42.1) |  |  |  |  |
| **Presence of parents<sup>VI</sup>** |  |  |  |  |  |  |
| Both present | 13,672 (44.7) | 2,693 (19.7) | 1 | <0.001 | 1 | <0.001 |
| Father absent | 15,471 (50.6) | 3,644 (23.6) | 1.33 (1.24–1.43) |  | 1.35 (1.25–1.46) |  |
| Mom absent | 178 (0.6) | 63 (35.4) | 2.23 (1.42–3.49) |  | 2.14 (1.34–3.44) |  |
| Neither present | 1,240 (4.1) | 486 (39.2) | 2.98 (2.45–3.62) |  | 2.98 (2.43–3.66) |  |
| **Wealth quintile** |  |  |  |  |  |  |
| 1st quintile | 2,828 (13.2) | 626 (22.1) | 1 | 0.669 |  |  |
| 2nd quintile | 3,208 (15.0) | 732 (22.8) | 1.07 (0.91–1.27) |  |  |  |
| 3rd quintile | 3,470 (16.2) | 759 (21.9) | 0.99 (0.84–1.16) |  |  |  |
| 4th quintile | 5,433 (25.4) | 1,170 (21.5) | 0.96 (0.83–1.12) |  |  |  |
| 5th quintile | 6,415 (30.0) | 1,488 (23.2) | 1.06 (0.92–1.23) |  |  |  |
| Missing | 9,207 (30.1) | 2,111 (22.93) |  |  |  |  |

<sup>I</sup> Children were defined as unvaccinated if they had not received any secondary series vaccinations (Measles and Yellow Fever) by 15 months of age.
<sup>II</sup> Controlled for sex, mothers age at birth, distance from health centre, presence of parents, immigration, headship, birth order and pregnancy type
<sup>III</sup> Controlled for sex, mother's age at birth, ethnicity, presence of parents, immigration and headship
<sup>IV</sup> Controlled for sex, mother's age at birth, ethnicity, distance from health centre, presence of parents, headship, birth order and pregnancy type
<sup>V</sup> Controlled for sex, mother's age at birth, ethnicity and distance from health centre
<sup>VI</sup> Controlled for sex, mother's age at birth, ethnicity, distance from health centre, immigration, birth order and pregnancy type
<sup>VII</sup> P-values obtained using Wald test

**Table 2A.**
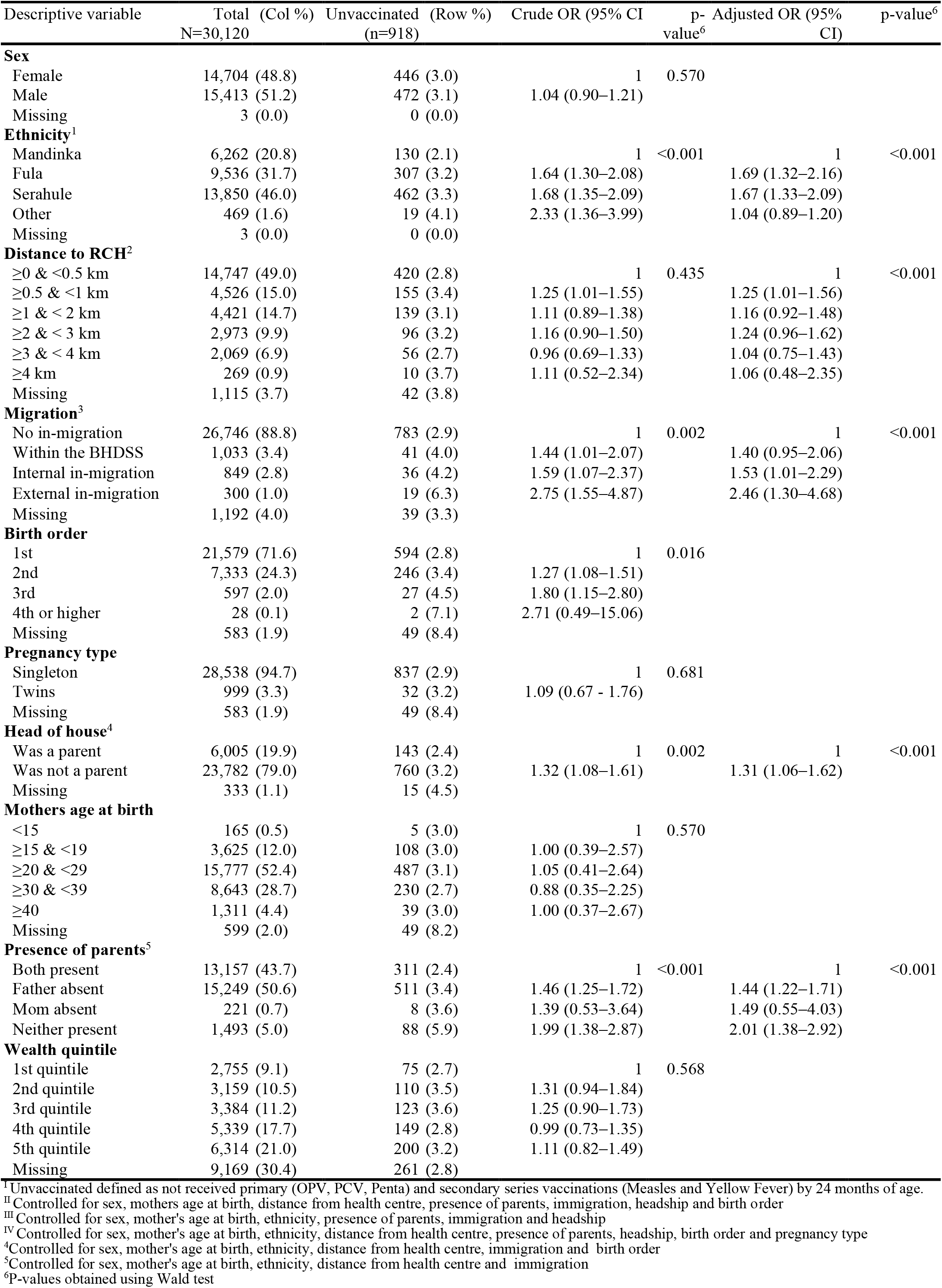
**Descriptive analysis of characteristics of children within the BHDSS and the crude and adjusted odds of being unvaccinated with the secondary vaccination series at 24-months of age.**

#### Multivariate secondary analysis

The results for the multivariate analysis, adjusting for potential confounders and clustering, for the secondary outcomes, vaccination status at 15- and 24-months of age are found in table 1A and table 2A. There remained strong evidence (p<0.001) that Mandinka children had lower odds of being unvaccinated compared to Fula and Serahule children at 15- and 24-months of age. In the adjusted models, there was some evidence that at 15- and 24-months of age, children who lived ≥0.5 and <1 km from a RCH had 12% and 25% higher odds of having missed their secondary series of vaccination compared to the children who lived <0.5 km (aOR 1.12, 95% CI 1.00–1.26 and aOR 1.25, 95% CI 1.01–1.55, respectively). There was no evidence for a difference in odds of being unvaccinated for children residing in any further distance parameters when compared to the nearest distance at both age points of interest (table 1A and table 2A). There was strong evidence (p<0.001) at both age points of interest for higher odds of being unvaccinated if there was only a single mother or neither parent present compared to children whose parents were both present. At 15- and 24- months of age the greatest increase in odds of being unvaccinated when compared to a child whose parents were present was if neither parent were present, (aOR 2.98, 95% CI 2.45–3.62 and aOR 1.99, 95% CI 1.38–2.87, respectively). There was evidence for higher odds of being unvaccinated if a child experienced external migration compared to children who had not experienced any immigration, at age 15- and 24-months (aOR 1.75, 95% CI 1.12–2.59 and aOR 2.46, 95% CI 1.30– 4.68, respectively). There were strong signs of negative confounding between model 1 and 2 for external migration at age 24-months as the point estimate increased by nearly 20% after adjusting for confounders (table 2A). There was no evidence of a difference in odds of non-vaccination for children at age 15-months who had internally migrated compared to children who had not immigrated (aOR 1.19, 95% CI 0.93–1.51) as the CI’s included unity, however at 24-months of age there was evidence of a difference in odds between these parameters (aOR 1.53, 95% CI 1.01–2.29). After controlling for confounders there was some evidence of increased odds for being unvaccinated at age 24-months in children whose parent were not the head of house compared to children whose parents were (aOR 1.31, 95% 1.06–1.62).

#### Death analysis

The death analysis, using Cox proportional hazards models satisfied the proportional hazards assumption through graphical assessments (figure 1A & figure 2A) and Schoenfeld residuals (p=0.722 and p=0.748, respectively).

**Figure 1A.**
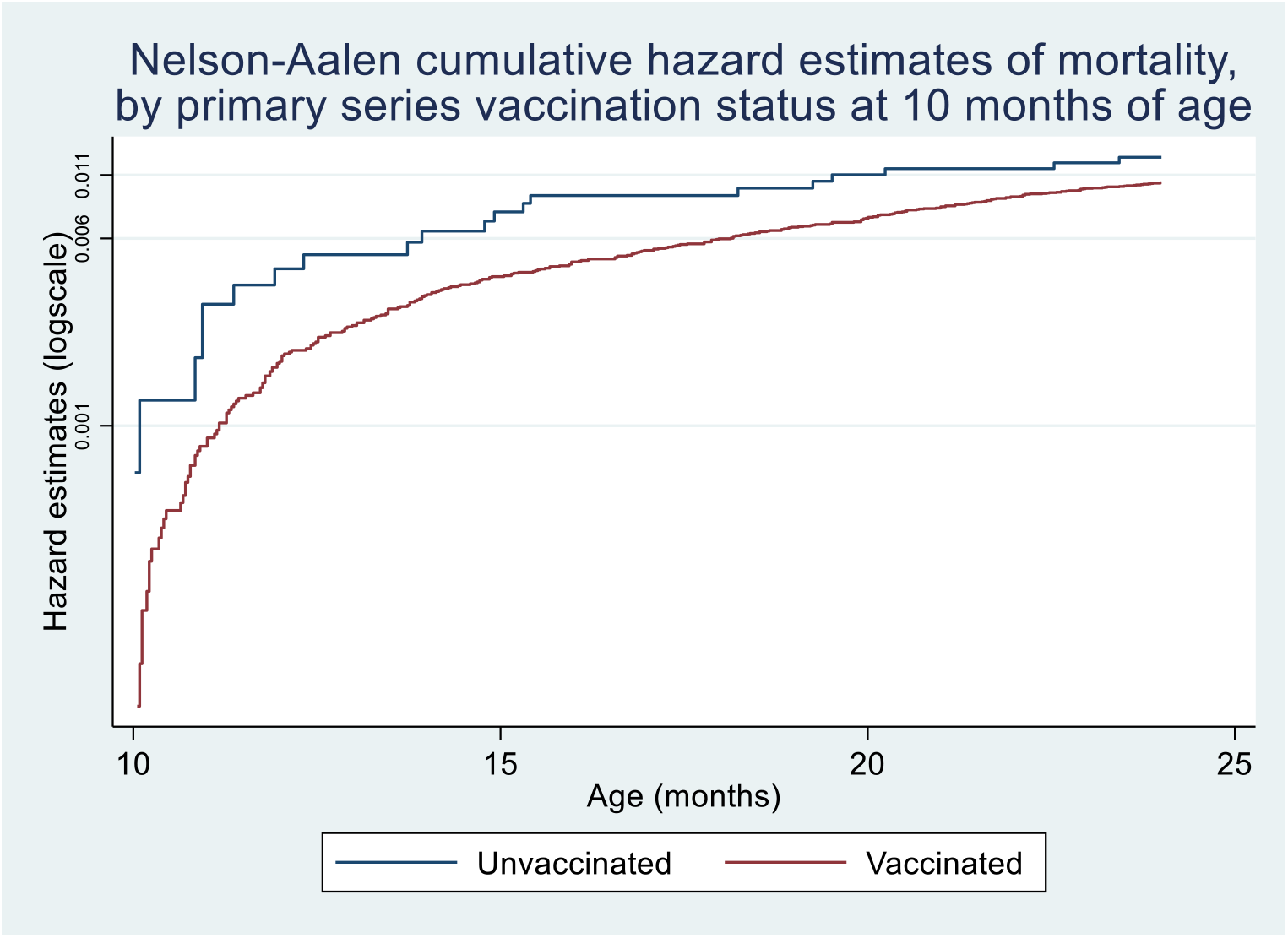
Survival analysis using Nelson-Aalen cumulative hazard estimates on the incidence of all-cause mortality between 10- and 24-months of age dependent on the primary series vaccination status at 10-months of age for children in The Gambia. Children were considered unvaccinated if they missed all their doses of the primary series.

**Figure 2A.**
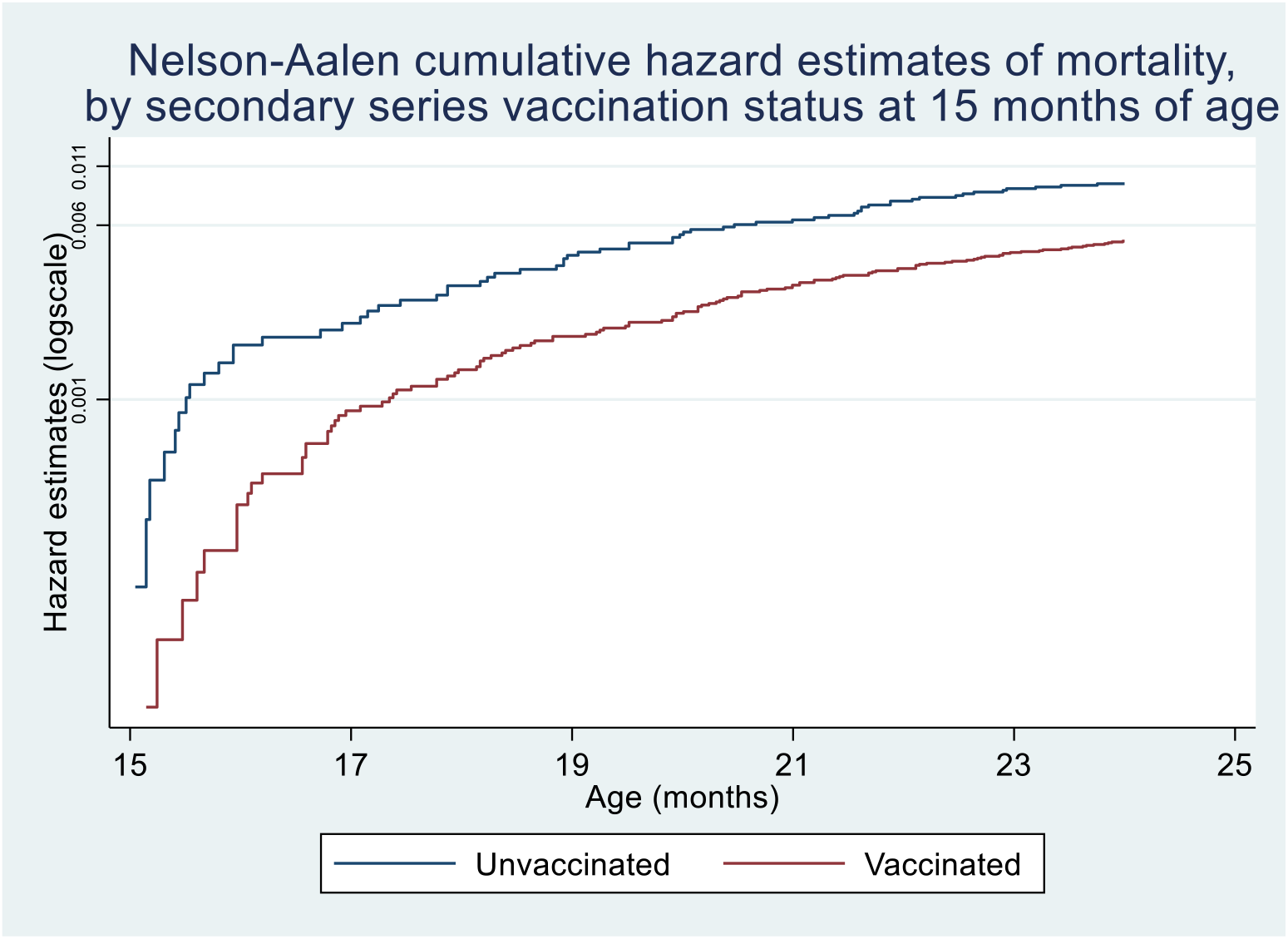
Survival analysis using Nelson-Aalen cumulative hazard estimates on the incidence of all-cause mortality between 15- and 24-months of age dependent on the secondary series vaccination status at 15-months of age for children in The Gambia. Children were considered unvaccinated if they missed all their doses of the secondary series.

